# Gene Signature for Predicting Metastasis in Prostate Cancer Using Primary Tumor Expression Profiles

**DOI:** 10.1101/2024.08.30.24312735

**Authors:** Itzel Valencia, Pier Vitale Nuzzo, Edoardo Francini, Francesco Ravera, Giuseppe Nicolò Fanelli, Sara Bleve, Cristian Scatena, Luigi Marchionni, Mohamed Omar

**Affiliations:** Department of Pathology and Laboratory Medicine, Weill Cornell Medicine, New York 10021 NY, USA; Dana-Farber Cancer Institute, Harvard University, Boston 02215 MA, USA; Department of Experimental and Clinical Medicine, University of Florence, 50134 Florence, Italy; Department of Internal Medicine and Medical Specialties, University of Genoa, 16132 Genoa, Italy; Department of Translational Medicine and New Technologies in Medicine and Surgery, University of Pisa, 56125 Pisa, Italy; Department of Medical Oncology, IRCCS Istituto Romagnolo per lo Studio dei Tumori (IRST) “Dino Amadori”, 47014 Meldola, Italy

## Abstract

Prostate cancer (PCa) is currently the most commonly diagnosed cancer and second leading cause of cancer-related death in men in the United States. The development of metastases is associated with a poor prognosis in PCa patients. Since current clinicopathological classification schemes are unable to accurately prognosticate the risk of metastasis for those diagnosed with localized PCa, there is a pressing need for precise and easily attainable biomarkers of metastatic risk in these patients. Primary tumor samples from 1239 individuals with PCa were divided into development (n=1000) and validation (n=239) cohorts. In the development cohort, we utilized a meta-analysis workflow on retrospective primary tumor gene expression profiles to identify a subset of genes predictive of metastasis. For each gene, we computed Hedges’ g effect size and combined their p-values using Fisher’s combined probability test. We then adjusted for multiple hypothesis testing using the Benjamini-Hochberg method. Our developed gene signature, termed Meta-Score, achieved a robust performance at predicting metastasis from primary tumor gene expression profiles, with an AUC of 0.72 in the validation cohort. In addition to its robust predictive power, Meta-Score also demonstrated a significant prognostic utility in two independent cohorts. Specifically, patients with a higher risk-score had a significantly worse metastasis-free survival and progression-free survival compared to those with lower score. Multivariate cox proportional hazards model showed that Meta-Score is significantly associated with worse survival even after adjusting for Gleason score. Our findings suggest that our primary tumor transcriptional signature, Meta-Score, could be a valuable tool to assess the risk of metastasis in PCa patients with localized disease, pending validation in large prospective studies.

**Author Summary:** Metastasis is the leading cause of death in patients diagnosed with prostate cancer (PCa), underscoring the need for reliable prediction tools to forecast the risk of metastasis at an early stage. Here, we utilize the gene expression profiles of 1,000 unique primary tumors from patients with localized PCa to develop a gene signature capable of predicting metastasis. Our signature, termed Meta-Score, comprises forty-five genes that can accurately distinguish primary tumor with high propensity for metastasis across different patient cohorts. Notably, Meta-Score maintained its robust predictive performance in an internal validation cohort of comprising primary tumor samples from 239 patients. In addition to its robust predictive performance, Meta-Score demonstrates a significant association with survival, independent of Gleason score in two independent patient cohorts, underscoring its prognostic utility. Taken together, Meta-Score is a robust risk-stratification tool that can be leveraged to identify patients at high-risk of metastasis and unfavorable survival using their primary tumor gene expression profiles.

## Introduction

Prostate cancer (PCa) is the most frequent non-cutaneous cancer and accounts for approximately 35,000 deaths every year in adult men in the United States (1). Localized PCa generally exhibits an indolent disease course, whereas de-novo metastatic PCa is associated with significant reductions in both overall survival and health-related quality of life, particularly after development of resistance to androgen deprivation therapy-based combinations (2).

Currently, clinical and pathological variables such as serum prostate-specific antigen (PSA) levels, PSA doubling time, TNM stage, and Gleason score, as well as genomic models, are routinely used to stratify localized PCa patients by their risk of metastatic disease and are used to decide their treatment (2,3). However, these classifications often fail to predict occurrence of metastatic disease as they do not fully capture PCa complexity and heterogeneity. Indeed, even apparently indolent tumors have metastatic potential while not all high-risk tumors develop metastases, resulting in a significant risk of under- or over-treatment, respectively, thus making the clinical management of PCa challenging. Therefore, there is an unmet clinical need for accurate and robust biomarkers of metastatic disease risk for patients with localized PCa (4).

To improve our understanding of the intricate mechanisms underlying the development of metastatic disease, ongoing research is exploring the genetic composition of PCa. PCa exhibits heterogeneous genetic profiles, characterized by the accumulation of somatic mutations, copy number alterations (CNAs), and chromosomal rearrangements. Common somatic alterations in PCa include *AR, TP53, EGFR, PTEN, RB1*, and homologous recombination repair genes, especially *BRCA1* and *BRCA2* (5–8). Despite the advances in understanding the genetic aberrations within PCa tumors, these factors alone are insufficient to fully predict the risk of metastasis of PCa. RNA sequencing (RNA-seq) provides a comprehensive evaluation of the transcriptional programs of PCa and its microenvironment (9,10). Over recent years, several studies have explored PCa gene expression profiling as a potential source of gene signatures associated with PCa outcomes (11–17). However, these studies are often characterized by a limited sample size, statistical power, and a short clinical follow-up, preventing the design of prospective trials necessary for the effective integration of transcriptomic gene signatures in the current clinical management of PCa.

Building on these results, our study aimed to develop and validate a robust transcriptomic gene signature for predicting PCa metastasis using primary tumor samples from patients with localized disease. To achieve this, we used retrospective primary tumor transcriptomic profiles of patients with localized PCa to develop a robust gene signature, Meta-Score, which was subsequently validated in a large internal patient cohort. This signature demonstrates high accuracy in predicting PCa metastasis and is significantly associated with metastasis-free survival (MFS) and progression-free survival (PFS) in two independent cohorts of patients with localized PCa. Overall, our results underscore the potential of transcriptomic gene signatures in enhancing the clinical management of localized and advanced PCa and pave the way for its evaluation in prospective clinical trials.

## Results

### Patient Characteristics

To develop the gene signature, we utilized the gene expression profiles of six different transcriptomics datasets, referred to as the development cohort, which comprises 1,000 primary tumor samples from PCa patients. Of these patients, 328 developed metastatic disease while 672 did not. To validate the predictive and prognostic performance of the gene signature, we used a large internal cohort (Nat. History), comprising 239 patients, of which 93 developed metastatic disease while 146 did not, with a median OS of 96 months and a median MFS of 84 months. Additionally, we examined the signature’s prognostic power in the TCGA PRAD cohort comprising 493 patients with a median PFS of 25.7 months, and a median OS of 30.3 months. Additional available clinical characteristics of these patients are shown in **Table S1**.

### Development of the Meta-Score Gene Signature Reveals Forty-five Putative Genes Related to PCa metastasis

We trained the meta-analysis algorithm to select genes that would be predictive of metastasis in primary PCa samples across the different six datasets comprising the training cohort. Through this method, we identified Meta-Score, which comprises 45 genes associated with the development of metastatic disease. Of these, 27 were upregulated in patients who developed metastasis, while 18 were downregulated. Additionally, the top five upregulated genes include *TMSB10, ENSA, ASPN, YWHAZ*, and *HES6,* while the top five down regulated genes include *KCTD14*, *AZGP1*, PART1, *CHRNA2*, and *DPT*. The full list of Meta-Score genes is provided in **Table S2.**

### Meta-Score Achieves a Robust Performance in Predicting Metastasis

In the pooled development cohort, comprising 1000 patients, Meta-Score achieved a pooled AUC of 0.81 (95% CI: 0.69-0.92I) (**Figure 1A**). In the internal validation cohort (Nat. History), Meta-Score maintained its robust predictive performance achieving an AUC of 0.72 (95% CI: 0.65-0.78), a balanced accuracy of 68%, and an MCC of 0.35 (**Figure 1B**).

**Fig 1.**
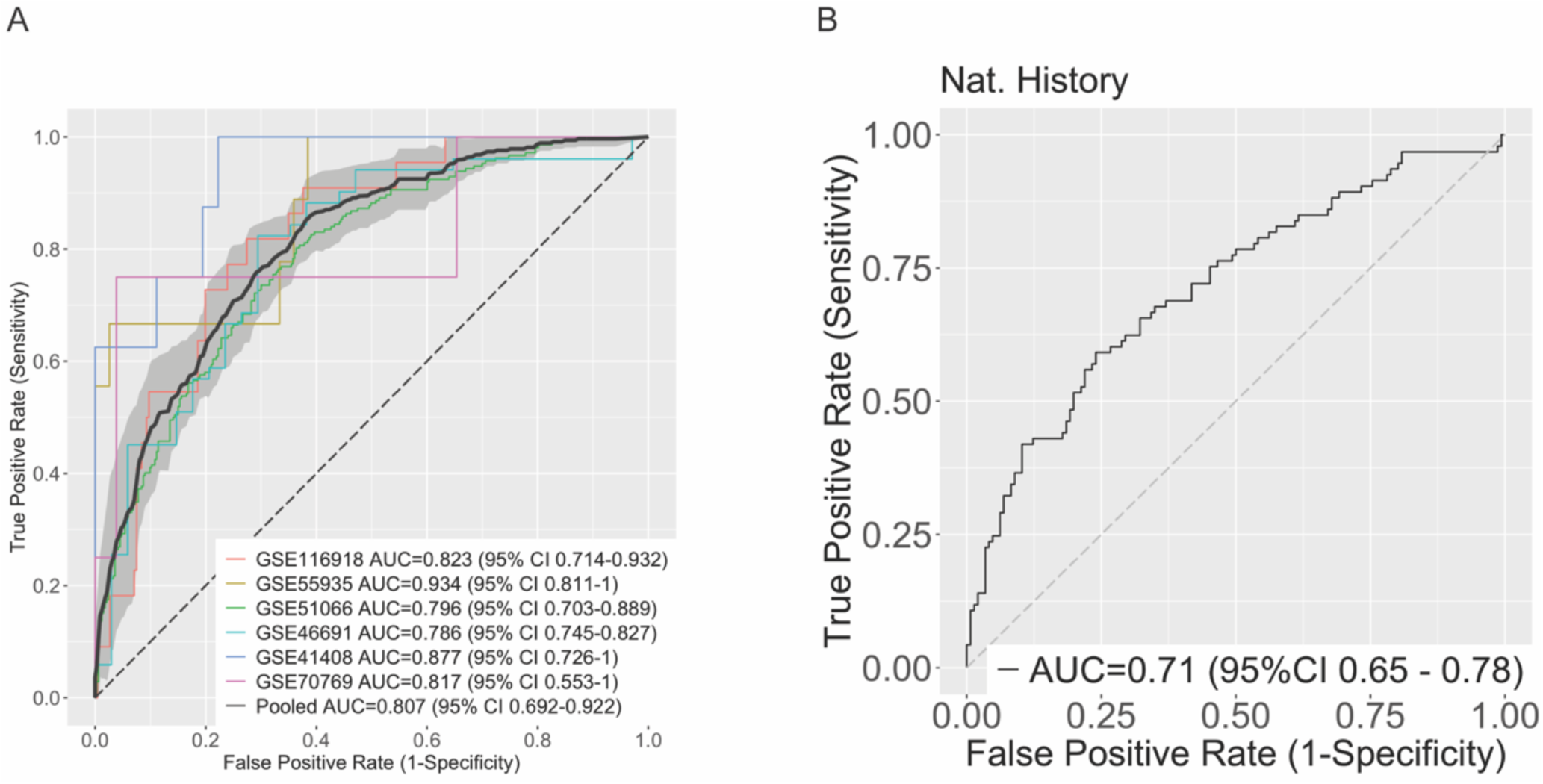
Development and validation of Meta-Score performance at predicting metastatic progression in patients with prostate cancer **A)** Receiver operating characteristics (ROC) curves and area under the ROC curve (AUC) showing the performance of Meta-Score at predicting metastasis in the pooled development cohort (n=1000 patients), comprising six different datasets. **B)** ROC curve and AUC of Meta-Score performance in the validation cohort (Nat. History) comprising 239 patients.

### Meta-Score is Significantly Associated with Survival in Two Independent Cohorts

To assess the prognostic utility of Meta-Score, we examined its association with OS, MFS, and PFS in the Nat. History and TCGA PRAD cohorts using univariate and multivariate survival analyses. Notably, when applying Meta-Score to the TCGA cohort, two upregulated (*ARL6IP1, SEM1*) and two downregulated (*KIAA1210, TMEM121B*) genes were not present in the expression profile of that cohort. In the Nat. History cohort, we found that high-risk patients, based on Meta-Score’s prediction, have a significantly shorter OS compared to low-risk patients in univariate survival analysis (logrank p-value = 0.013) (**Figure 2A**), but its significance is lost after adjusting for Gleason score (HR=1.6, 95% CI: 0.81-3.2) (**Figure 2B**). Additionally, high-risk patients were found to have a significantly shorter MFS compared to low-risk patients (logrank p-value < 0.0001) (**Figure 2C**), even after adjusting for Gleason score (HR= 2.0, 95% Cl: 1.2-3.2) (**Figure 2D**). These results were further confirmed in the TCGA PRAD cohort, in which high-risk patients had a significantly shorter PFS than low-risk patients (logrank p-value < 0.0001) (**Figure 2E**), even after adjusting for Gleason score (HR=1.6, 95% CI: 1.04-2.6) (**Figure 2F**). These results underscore the prognostic utility of Meta-Score and its potential to provide valuable insights into improving the risk stratification of patients affected with PCa.

**Fig 2.**
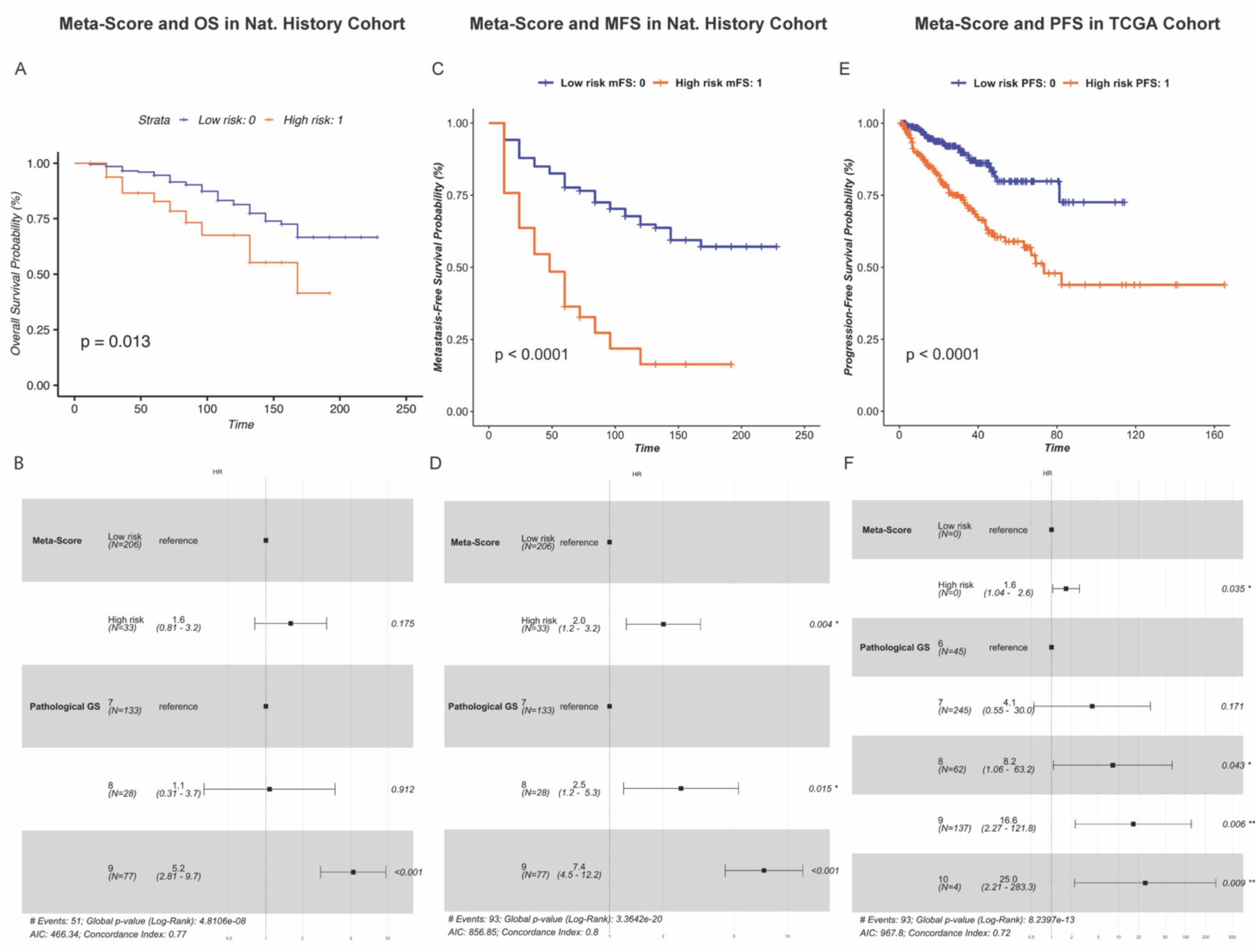
Meta-Score is significantly associated with patients’ survival in the Nat. History and TCGA PRAD cohorts **A-B**) Meta-Score and overall survival (OS) in the Nat. History cohort using univariate and multivariate survival analysis. The Kaplan-Meier (KM) chart (**A**) shows the difference in survival between patients predicted as high- and low-risk based on Meta-Score. P: logrank p-value. The forest plot (**B**) displays the hazard ratios (HR) and 95% confidence intervals (CI) of Meta-Score and Gleason score resulting from multivariate Cox proportional hazards (CPH) model. P: Wald test p-value. **C-D**) Meta-Score and metastasis-free survival (MFS) in the Nat. History Cohort. Survival analysis was performed using KM survival estimates (**C**) and multivariate survival, adjusting for Gleason score using CPH model (**D**). **E-F**) Meta-Score and progression-free survival (PFS) in the TCGA PRAD cohort using univariate (**E**) and multivariate (**F**) survival analysis.

### Meta-Score Outperforms Decipher in Predicting Clinical Outcomes of PCa Patients

Decipher is a genomic test that has been proposed to assist physicians in their decision on treatment options for localized PCa. To compare Meta-Score to Decipher we used a logistic regression model to fit each signature to patients’ clinical outcome and compute risk-scores. In the Nat. History cohort, Decipher was not significantly associated with OS in both univariate (logrank p-value=0.23, (**Figure 3A**) and multivariate (HR=1.3, 95% CI: 0.72-2.3, **Figure 3B**) survival analyses. However, we found that Decipher is significantly associated with MFS (logrank p-value=0.0002, **Figure 3C**) even after adjusting for Gleason score (HR=1.8, 95% CI: 1.2-2.7, **Figure 3D**). In the TCGA PRAD cohort, while Decipher was found to be significantly associated with PFS in univariate analysis (logrank p-value<0.0001, **Figure 3E**), it lost its significance after adjusting for Gleason score (HR=1.0, 95% CI: 0.64-1.7, **Figure 3F**). Overall, these results show that Decipher does not outperform Meta-Score in either our internal validation cohort or in the TCGA PRAD cohort in terms of survival analyses.

**Fig 3.**
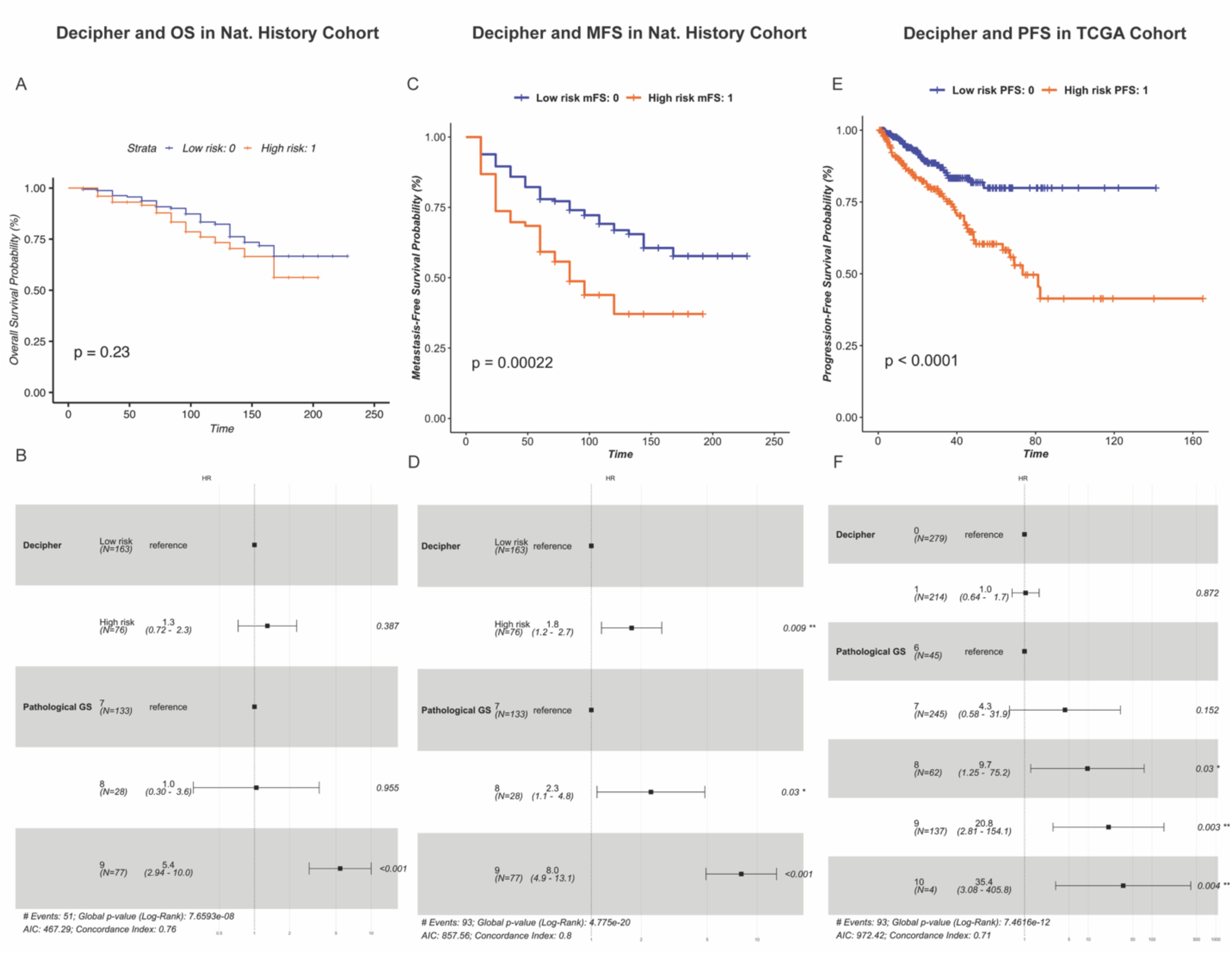
Prognostic performance of DECIPHER in the Nat. History and TCGA PRAD cohorts. **A-B**) Decipher and overall survival (OS) in the Nat. History cohort using univariate and multivariate survival analysis. The Kaplan-Meier (KM) chart (**A**) shows the difference in survival between patients predicted as high- and low-risk based on Decipher. P: logrank p-value. The forest plot (**B**) displays the hazard ratios (HR) and 95% confidence intervals (CI) of Decipher and Gleason score resulting from multivariate Cox proportional hazards (CPH) model. P: Wald test p- value. **C-D**) Decipher and metastasis-free survival (MFS) in the Nat. History Cohort. Survival analysis was performed using KM survival estimates (**C**) and multivariate survival, adjusting for Gleason score using CPH model (**D**). **E-F**) Decipher and progression-free survival (PFS) in the TCGA PRAD cohort using univariate (**E**) and multivariate (**F**) survival analysis.

## Discussion

Over the last decade, numerous studies have tried to address the clinical need for predicting aggressive PCa through the use of individual biomarkers or gene expression signatures (10,18–20). Despite these efforts, the clinical implementation of these predictive tools has been limited, primarily due to their inability to demonstrate significant improvements over established clinical parameters, such as Gleason score. The advent and rapid advancement of machine learning technologies has contributed to the evolution of research in the discovery and validation of cancer biomarkers, including ones for PCa. These advancements have facilitated the development of various predictive strategies, including the development of predictive gene expression signatures (21). However, despite these technological and methodological advancements, the clinical landscape still lacks a validated biomarker panel capable of reliably predicting metastasis in localized PCa.

Here, we introduce a novel gene signature, Meta-Score, which is capable of predicting metastatic events using the gene expression profiles of primary tumors derived from patients with localized PCa. This signature, comprising 27 up-regulated and 18 down-regulated genes, was discovered through a large-scale meta-analysis of transcriptomic profiles of primary tumor samples, derived from six different retrospective patient cohorts. Subsequently, we validated the signature’s predictive and prognostic performance in two independent patient cohorts, in which Meta-Score achieved a robust performance at differentiating primary prostate tumors likely to metastasize from those at low-risk of developing metastasis. Additionally, we found that Meta-Score is significantly associated with MFS and PFS even after adjusting for Gleason score, further underscoring its independent prognostic utility.

Current prognostic models for metastatic disease in localized PCa fail to reliably predict the occurrence of metastatic disease, leading to potential under- or overtreatment [54]. Presently, Decipher (22), a tissue-based genomic test, is used by physicians to make treatment decisions in patients presenting with localized PCa. Although Decipher is favored, here we demonstrate that our Meta-Score signature might be better at determining a more accurate risk assessment in regard to MFS and PFS. Importantly, Meta-Score incorporates the expression of genes involved in critical molecular pathways, enhancing the precision of existing risk stratification methods.

Notably, the genes comprising Meta-Score offer valuable insights into the biological mechanisms driving metastasis in PCa. Several of these genes are implicated in PCa-related signaling pathways. For instance, *ASPN* variants have been linked to the TGFβ signaling pathway [39]. In early prostate cancer, TGFβ inhibits tumor growth, but as the disease progresses, tumors often become resistant (23). Additionally, *HES6* has been shown to enhance transcriptional activity of *AR* and serve as a potential biomarker for neuroendocrine PCa (24). PCa cells harboring *TMPRSS-ERG* gene fusion can regulate *CXCR4* transcription (25). Activation of this receptor is implicated in various cancer related processes such as migration, adhesion and metastasis (26–28). Furthermore, *SOX4* may function as a transforming oncogene that may promote epithelial-mesenchymal transition (EMT) (29). Our signature also comprises genes that serve as molecular markers in the progression and severity of PCa. For example, *ASNS* upregulation has been observed in castration-resistant prostate cancer (CRPC), indicative of disease progression (30). Furthermore, *ALDH1A1* interaction with *AR* and retinoid receptor (RAR) transcriptional programs have been shown to be associated with PCa metastasis (31). Moreover, increased *YWHAZ* expression strongly correlates with a high Gleason score (>= 8) at diagnosis, and also with PSA relapse (32) . On the other hand, PCa cells have exhibited low levels of *KCTD11* mRNA (33). The precise roles of *STC2*, predicted to be a secreted glycosylated protein, and *RNF19*, encoding an E3 ubiquitin ligase, in promoting PCa cell growth (34) and their association with clinically higher expression and advanced Gleason score, respectively, remains unclear (35). Additionally, *AZGP1* acts as a negative regulator of angiogenesis; its loss promotes angiogenesis in prostate cancer (36).

One of the strengths of our study is the large sample size, encompassing 1239 primary PCa samples enhances the statistical power and robustness of the findings. The utilization of the MetaIntegrator R package and the meta-analysis workflow—computing Hedges effect sizes, combining p-values using Fisher’s exact method, and implementing Benjamini-Hochberg FDR correction—ensures a thorough and reliable identification of predictive genes. This multi-step approach minimizes the risk of false positives and enhances the credibility of the gene signature. Additionally, integrating RNA-seq data from six different cohorts, provides a detailed evaluation of transcriptional programs associated with metastasis in PCa. This large-scale, multi-cohort analysis addresses the limitations of previous studies that were constrained by smaller sample sizes and less diverse datasets. Furthermore, the study demonstrates the clinical relevance of Meta-Score by showing its association with OS, MFS, and PFS, independent of Gleason score. Despite its strengths, this study has some inherent limitations. The retrospective nature of the analysis introduces potential biases related to patient selection and data quality. The heterogeneity of the datasets is another limitation. Variations in sample processing, sequencing platforms, and data collection methods across cohorts could introduce biases. These technical variations may impact the gene expression profiles and the accuracy of the identified gene signature. Notably, the original set of twenty-two genes from Decipher was narrowed down to fifteen, due to the absence of seven genes across the training, testing, and TCGA datasets. Furthermore, while the development dataset comprised a substantial sample size of over 1000, access to comprehensive patient information was limited. Specifically, details regarding ethnic backgrounds, age, race, and other demographic information were incomplete. Consequently, we cannot assert the diversity of our cohort, suggesting that the gene signature may not be applicable to a diverse population. Additionally, clinical variables such as stage and PSA levels pre-surgery were not available across all datasets. The lack of follow-up information in the training cohort also precluded the analysis of OS, MFS, and PFS, which would have provided additional validation of the gene signature’s prognostic value.

Nonetheless, our meta-analysis-derived signature offers a thorough and resilient analysis by amalgamating data from various studies, thereby enhancing the generalizability of the findings. By refining Meta-Score based on the weighted AUC, we prioritized both predictive accuracy and interpretational simplicity, potentially easing its clinical implementation. Further research and validation in larger, prospective clinical trials are essential to evaluate the practical utility of our Meta-Score signature in clinical settings, potentially enabling its integration alongside well-established nomograms. Such outcome could refine patient management strategies, allowing for personalized treatment plans that consider both traditional prognostic factors and molecular signatures.

In conclusion, Meta-Score represents a significant step forward in the prognostic assessment of metastasis risk in localized PCa. By leveraging gene expression profiles from primary tumors, this tool offers a valuable addition to existing clinical models. Future research is needed to validate these findings in prospective clinical studies paving the way for its integration into routine clinical practice.

## Methods

### Data acquisition

We performed a rigorous search of the Gene Expression Omnibus (GEO) database (37) using the search terms: prostate cancer AND Homo sapiens, entry type: series, study type: expression profiling by array. These terms were used to identify six gene expression profiling datasets derived from 1000 unique localized primary PCa samples which included information on whether a metastatic event occurred or not. Additionally, for independent validation of the signature performance, we used an internal cohort comprising 239 patients who underwent radical prostatectomy for localized PCa (henceforth termed Nat. History cohort) (**Table S1**). To further investigate the prognostic utility of the signature, we stratified patients into risk groups and examined their association with survival in the Nat. History cohort as well as the Cancer Genome Atlas (TCGA) prostate adenocarcinoma (PRAD) cohort, comprising 493 patients.

### Data Preprocessing and Quality Control

For each dataset, the gene expression profiles were extracted and filtered based on the coefficient of variation, using thresholds ranging from 0.1 to 10, followed by log2 transformation. Subsequently, we extracted the patient metadata from each dataset to link the expression profiles to clinical data. To maintain data cohesion, we filtered out patients with unknown metastasis status and kept those with binary metastasis indicator. Once the clinical and gene expression data was processed, we integrated the processed data by replacing the original datasets. Subsequently, the six training datasets were merged into one cohesive training metadata set based on a set of common genes (n=18,550 genes). The Nat. History cohort (n=239 patients) was processed using the same approaches implemented for the training data.

### Meta-Analysis of Primary PCa Gene Expression Data

To streamline our analysis of multiple datasets, we used a gene expression meta-analysis workflow described by Haynes *et al.* (38) and implemented within the R package *MetaIntegrator*. Briefly, this workflow computes a Hedges effect size (39) for each gene in each dataset. These effect sizes are then pooled across all the datasets using a random effect model by assuming that results from each study are drawn from a single distribution and that each inter-study difference is just a random effect (40). The approach computes the log sum of p-values that each gene is up/down-regulated, then combines p-values using Fisher’s exact method, and finally performs Benjamini-Hochberg false discovery rate (FDR) correction across all genes (41). To develop our gene signature, we deployed this algorithm on our metadata to identify which genes were able to forecast the occurrence of metastasis in the primary PCa specimens.

### Feature Selection

With the aim of developing a robust gene signature, we emphasized gene expression patterns as a feature selection for our metadata. In our analysis, we filtered genes by keeping only those with an effect size *>* 0.2, an FDR cut-off of 0.05, and present in at least four datasets. Additionally, to further optimize the gene signature, we performed a forward search process which allowed us to select for the most relevant genes, measured by the combined weighted Area Under the ROC curve (AUC), to accurately predict metastasis in the primary PCa samples, using the following formula:

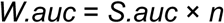

Where *W.auc* is the weighted AUC, *S.auc* is the sum of AUC of each data set and *n* is the number of samples in the data set. Each gene is iteratively assessed until the threshold is reached and only those that contribute the greatest weighted AUC are kept in the final signature.

### Software and Packages

All steps of this analysis utilized R version 4.3.3. The multi-cohort analysis of the gene expression data from the testing dataset was performed using the MetaIntegrator package (42).Additionally, the survival analysis was performed with the survival (43,44) and survminer (45) packages.

### Independent Assessment of the Signature Predictive Performance

The accuracy of our signature in predicting PCa metastasis was assessed in the independent validation cohort (n=239 patients) using various performance metrics including the area under the receiver operating characteristics curve (AUC-ROC), balanced accuracy, and Matthew’s correlation coefficient (MCC). The ROC curve and AUC were computed using the prediction probability scores (risk-scores) derived from the signature. Prediction labels for each patient were computed by categorizing the computed risk-score into binary labels based on the best threshold derived from the ROC curve of the training data. Other performance metrics, including balanced accuracy, sensitivity, specificity, and MCC were computed by comparing the prediction labels to the ground-truth patient labels.

### Prognostic evaluation of the Signature

To evaluate the prognostic utility of our gene signature, we computed its association with survival in the Nat. History cohort (n=239 patients) as well as the TCGA PRAD cohort (n=493 patients). Specifically, we compared the probability of overall survival (OS), metastasis-free survival (MFS), and progression-free survival (PFS), between low- and high-risk patients, identified by our signature, using Kaplan-Meier (KM) survival estimates log-rank test. To assess the independent prognostic value of Meta-Score, we used multivariate Cox proportional hazards (CPH) model and Wald test to compute the hazard ratio (HR) and 95% confidence intervals (CIs) of the signature’s risk-scores after adjusting for important clinical and pathological variables. Finally, we compared the prognostic utility of the signature to that of Decipher, a well-known genomic test used in clinical settings.

### Availability of Data and Materials

The expression profiles of the development cohort are derived from six publicly available datasets which are available through the Gene Expression Omnibus (GEO) under the accession codes GSE116918, GSE55935, GSE51066, GSE46691, GSE41408, and GSE70769. The Natural History Cohort is an internal validation cohort and will be provided upon request. The TCGA Prostate Adenocarcinoma dataset used in this study is publicly available through cBioPortal using the following links: https://www.cbioportal.org/study/summary?id=prad_tcga_pan_can_atlas_2018.

### Code Reporting

The code used to perform the analysis can be accessed using the following GitHub repository https://github.com/itzvals/Met_PCa_signature

## Data Availability

All data produced in the present study are available upon reasonable request to the authors.

https://www.cbioportal.org/study/summary?id=prad_tcga_pan_can_atlas_2018

https://github.com/itzvals/Met_PCa_signature

## Acknowledgments

This work was supported by the National Cancer Institute grant R01 CA200859 (to L.M.)

## Author Contributions

M.O. formulated the research question and supervised the study. I.V. performed the analysis. I.V., P.V.N., E.F., F.R., G.N.F., S.B., and C.S. wrote the manuscript draft. E.F., F.R., G.N.F., S.B., C.S., L.M., and M.O. edited the manuscript. All authors have read and agreed to the final version of the manuscript.

## Supplemental table titles and legends

**Table S1. Clinical features of the datasets used in this analysis** The development cohorts were used to train Meta-Score and the validation cohort was used to assess its performance. To inspect Meta-Scores impact on survival both the validation and TCGA PRAD cohort were utilized.

**Table S2. List of upregulated and downregulated genes in Meta-Score** The signature is comprised of 27 upregulated and 18 down regulated genes. For each gene we provide a summary of the respective criteria that was used to construct the gene signature, including effect size, p-value and adjusted p-value.

## References

1. Siegel RL, Miller KD, Wagle NS, Jemal A. Cancer statistics, 2023. CA Cancer J Clin. 2023;73(1):17–48.

2. Parker C, Castro E, Fizazi K, Heidenreich A, Ost P, Procopio G, et al. Prostate cancer: ESMO Clinical Practice Guidelines for diagnosis, treatment and follow-up. Ann Oncol. 2020 Sep;31(9):1119–34.

3. Jenkins V, Solis-Trapala I, Payne H, Mason M, Fallowfield L, May S, et al. Treatment Experiences, Information Needs, Pain and Quality of Life in Men with Metastatic Castrate-resistant Prostate Cancer: Results from the EXTREQOL Study. Clin Oncol. 2019 Feb;31(2):99–107.

4. Sartor O, Eisenberger M, Kattan MW, Tombal B, Lecouvet F. Unmet needs in the prediction and detection of metastases in prostate cancer. The Oncologist. 2013;18(5):549–57.

5. Spisak S, Tisza V, Nuzzo PV, Seo JH, Pataki B, Ribli D, et al. A biallelic multiple nucleotide length polymorphism explains functional causality at 5p15.33 prostate cancer risk locus. Nat Commun. 2023 Aug 23;14(1):5118.

6. Conteduca V, Oromendia C, Eng KW, Bareja R, Sigouros M, Molina A, et al. Clinical features of neuroendocrine prostate cancer. Eur J Cancer Oxf Engl 1990. 2019 Nov;121:7–18.

7. De Muga S, Hernández S, Agell L, Salido M, Juanpere N, Lorenzo M, et al. Molecular alterations of EGFR and PTEN in prostate cancer: association with high-grade and advanced-stage carcinomas. Mod Pathol. 2010 May;23(5):703–12.

8. Shah S, Rachmat R, Enyioma S, Ghose A, Revythis A, Boussios S. BRCA Mutations in Prostate Cancer: Assessment, Implications and Treatment Considerations. Int J Mol Sci. 2021 Nov 23;22(23):12628.

9. Schmidt L, Møller M, Haldrup C, Strand SH, Vang S, Hedegaard J, et al. Exploring the transcriptome of hormone-naive multifocal prostate cancer and matched lymph node metastases. Br J Cancer. 2018 Dec;119(12):1527–37.

10. Ma C, Zhou Y, Fanelli GN, Stopsack KH, Fiorentino M, Zadra G, et al. The Prostate Stromal Transcriptome in Aggressive and Lethal Prostate Cancer. Mol Cancer Res. 2023 Mar 1;21(3):253–60.

11. Yimamu Y, Yang X, Chen J, Luo C, Xiao W, Guan H, et al. The Development of a Gleason Score-Related Gene Signature for Predicting the Prognosis of Prostate Cancer. J Clin Med. 2022 Dec 1;11(23):7164.

12. Yang L, Roberts D, Takhar M, Erho N, Bibby BAS, Thiruthaneeswaran N, et al. Development and Validation of a 28-gene Hypoxia-related Prognostic Signature for Localized Prostate Cancer. EBioMedicine. 2018 May;31:182–9.

13. Mohammad T, Singh P, Jairajpuri DS, Al-Keridis LA, Alshammari N, Adnan Mohd, et al. Differential Gene Expression and Weighted Correlation Network Dynamics in High-Throughput Datasets of Prostate Cancer. Front Oncol. 2022 Jun 1;12:881246.

14. Pakula H, Omar M, Carelli R, Pederzoli F, Fanelli GN, Pannellini T, et al. Distinct mesenchymal cell states mediate prostate cancer progression. Nat Commun. 2024 Jan 8;15(1):363.

15. Davidson SM, Schmidt DR, Heyman JE, O’Brien JP, Liu AC, Israelsen WJ, et al. Pyruvate Kinase M1 Suppresses Development and Progression of Prostate Adenocarcinoma. Cancer Res. 2022 Jul 5;82(13):2403–16.

16. Erho N, Crisan A, Vergara IA, Mitra AP, Ghadessi M, Buerki C, et al. Discovery and Validation of a Prostate Cancer Genomic Classifier that Predicts Early Metastasis Following Radical Prostatectomy. Creighton C, editor. PLoS ONE. 2013 Jun 24;8(6):e66855.

17. Zhao SG, Evans JR, Kothari V, Sun G, Larm A, Mondine V, et al. The Landscape of Prognostic Outlier Genes in High-Risk Prostate Cancer. Clin Cancer Res. 2016 Apr 1;22(7):1777–86.

18. Penney KL, Tyekucheva S, Rosenthal J, El Fandy H, Carelli R, Borgstein S, et al. Metabolomics of Prostate Cancer Gleason Score in Tumor Tissue and Serum. Mol Cancer Res. 2021 Mar 1;19(3):475–84.

19. Nuzzo PV, Ravera F, Saieva C, Zanardi E, Fotia G, Malgeri A, et al. Clinical outcomes of volume of disease on patients receiving enzalutamide *versus* abiraterone acetate plus prednisone as first-line therapy for metastatic castration-resistant prostate cancer. Ther Adv Med Oncol. 2023 Jan;15:17588359231156147.

20. Nuzzo PV, Pederzoli F, Saieva C, Zanardi E, Fotia G, Malgeri A, et al. Clinical impact of volume of disease and time of metastatic disease presentation on patients receiving enzalutamide or abiraterone acetate plus prednisone as first-line therapy for metastatic castration-resistant prostate cancer. J Transl Med. 2023 Feb 3;21(1):75.

21. Omar M, Nuzzo PV, Ravera F, Bleve S, Fanelli GN, Zanettini C, et al. Notch-based gene signature for predicting the response to neoadjuvant chemotherapy in triple-negative breast cancer. J Transl Med. 2023 Nov 15;21(1):811.

22. Spratt DE, Yousefi K, Deheshi S, Ross AE, Den RB, Schaeffer EM, et al. Individual Patient-Level Meta-Analysis of the Performance of the Decipher Genomic Classifier in High-Risk Men After Prostatectomy to Predict Development of Metastatic Disease. J Clin Oncol. 2017 Jun 20;35(18):1991–8.

23. Thompson-Elliott B, Johnson R, Khan SA. Alterations in TGFβ signaling during prostate cancer progression. Am J Clin Exp Urol. 2021;9(4):318–28.

24. Vias M, Massie CE, East P, Scott H, Warren A, Zhou Z, et al. Pro-neural transcription factors as cancer markers. BMC Med Genomics. 2008 May 19;1:17.

25. Singareddy R, Semaan L, Conley-LaComb MK, St. John J, Powell K, Iyer M, et al. Transcriptional Regulation of CXCR4 in Prostate Cancer: Significance of TMPRSS2-ERG Fusions. Mol Cancer Res. 2013 Nov 1;11(11):1349–61.

26. Kukreja P, Abdel-Mageed AB, Mondal D, Liu K, Agrawal KC. Up-regulation of CXCR4 Expression in PC-3 Cells by Stromal-Derived Factor-1α (CXCL12) Increases Endothelial Adhesion and Transendothelial Migration: Role of MEK/ERK Signaling Pathway–Dependent NF-κB Activation. Cancer Res. 2005 Nov 1;65(21):9891–8.

27. Engl T, Relja B, Marian D, Blumenberg C, Müller I, Beecken WD, et al. CXCR4 Chemokine Receptor Mediates Prostate Tumor Cell Adhesion through α5 and β3 Integrins. Neoplasia. 2006 Apr;8(4):290–301.

28. Chinni SR, Sivalogan S, Dong Z, Filho JCT, Deng X, Bonfil RD, et al. CXCL12/CXCR4 signaling activates Akt-1 and MMP-9 expression in prostate cancer cells: The role of bone microenvironment-associated CXCL12. The Prostate. 2006 Jan;66(1):32–48.

29. Wang L, Zhang J, Yang X, Chang YWY, Qi M, Zhou Z, et al. SOX4 is associated with poor prognosis in prostate cancer and promotes epithelial–mesenchymal transition in vitro. Prostate Cancer Prostatic Dis. 2013 Dec;16(4):301–7.

30. Sircar K, Huang H, Hu L, Cogdell D, Dhillon J, Tzelepi V, et al. Integrative molecular profiling reveals asparagine synthetase is a target in castration-resistant prostate cancer. Am J Pathol. 2012 Mar;180(3):895–903.

31. Gorodetska I, Offermann A, Püschel J, Lukiyanchuk V, Gaete D, Kurzyukova A, et al. ALDH1A1 drives prostate cancer metastases and radioresistance by interplay with AR- and RAR-dependent transcription. Theranostics. 2024;14(2):714–37.

32. Rüenauver K, Menon R, Svensson MA, Carlsson J, Vogel W, Andrén O, et al. Prognostic significance of YWHAZ expression in localized prostate cancer. Prostate Cancer Prostatic Dis. 2014 Dec;17(4):310–4.

33. Zazzeroni F, Nicosia D, Tessitore A, Gallo R, Verzella D, Fischietti M, et al. *KCTD11* Tumor Suppressor Gene Expression Is Reduced in Prostate Adenocarcinoma. BioMed Res Int. 2014;2014:1–9.

34. Tamura K, Furihata M, Chung S, Uemura M, Yoshioka H, Iiyama T, et al. Stanniocalcin 2 overexpression in castration-resistant prostate cancer and aggressive prostate cancer. Cancer Sci. 2009 May;100(5):914–9.

35. Zhang N, Huang D, Ruan X, Ng ATL, Tsu JHL, Jiang G, et al. CRISPR screening reveals gleason score and castration resistance related oncodriver ring finger protein 19 A (RNF19A) in prostate cancer. Drug Resist Updat. 2023 Mar;67:100912.

36. Wen RM, Qiu Z, Marti GEW, Peterson EE, Marques FJG, Bermudez A, et al. AZGP1 deficiency promotes angiogenesis in prostate cancer. J Transl Med. 2024 Apr 24;22(1):383.

37. Barrett T, Wilhite SE, Ledoux P, Evangelista C, Kim IF, Tomashevsky M, et al. NCBI GEO: archive for functional genomics data sets—update. Nucleic Acids Res. 2012 Nov 26;41(D1):D991–5.

38. Haynes WA, Vallania F, Liu C, Bongen E, Tomczak A, Andres-Terrè M, et al. EMPOWERING MULTI-COHORT GENE EXPRESSION ANALYSIS TO INCREASE REPRODUCIBILITY. Pac Symp Biocomput Pac Symp Biocomput. 2017;22:144–53.

39. Hedges LV. Distribution Theory for Glass’s Estimator of Effect size and Related Estimators. J Educ Stat. 1981 Jun;6(2):107–28.

40. DerSimonian R, Kacker R. Random-effects model for meta-analysis of clinical trials: An update. Contemp Clin Trials. 2007 Feb;28(2):105–14.

41. Benjamini Y, Hochberg Y. Controlling the False Discovery Rate: A Practical and Powerful Approach to Multiple Testing. J R Stat Soc Ser B Stat Methodol. 1995 Jan 1;57(1):289–300.

42. Winston A. Haynes, Francesco Vallania, Aurelie Tomczak, Timothy Sweeney, Erika Bongen, Aditya M. Rao, Purvesh Khatri. MetaIntegrator: Meta-Analysis of Gene Expression Data [Internet]. 2016 [cited 2024 Jul 25]. p. 2.1.3. Available from: https://CRAN.R-project.org/package=MetaIntegrator

43. Therneau TM. survival: Survival Analysis [Internet]. 2001 [cited 2024 Jul 25]. p. 3.7-0. Available from: https://CRAN.R-project.org/package=survival

44. Therneau TM, Grambsch PM. The Cox Model. In: Modeling Survival Data: Extending the Cox Model [Internet]. New York, NY: Springer New York; 2000 [cited 2024 Jul 25]. p. 39–77. (Dietz K, Gail M, Krickeberg K, Samet J, Tsiatis A, editors. Statistics for Biology and Health). Available from: http://link.springer.com/10.1007/978-1-4757-3294-8_3

45. Kassambara A, Kosinski M, Biecek P. survminer: Drawing Survival Curves using “ggplot2” [Internet]. 2016 [cited 2024 Jul 25]. p. 0.4.9. Available from: https://CRAN.R-project.org/package=survminer

