## Supplementary material for "Gene Signature for Predicting Metastasis in Prostate Cancer Using Primary Tumor Expression Profiles": Table S1

| | DEVELOPMENT COHORTS* | | | | | | | VALIDATION COHORT # | TCGA PRAD COHORT§ |
|---|---|---|---|---|---|---|---|---|---|
| | GSE116918 | GSE55935 | GSE51066 | GSE46691 | GSE41408 | GSE70769 | Total | | |
| **Patients** | 248 | 44 | 85 | 545 | 48 | 30 | 1000 | 239 | 493 |
| **Pathological Stage** | | | | | | | | | |
| T1 | 51 | - | - | - | 0 | 14 | 65 | 101 | 0 |
| T2 | 76 | - | - | - | 15 | 11 | 102 | 130 | 186 |
| T3 | 92 | - | - | - | 26 | 4 | 122 | 7 | 290 |
| T4 | 4 | - | - | - | 6 | 0 | 10 | 0 | 10 |
| *NA* | 25 | 44 | 85 | 545 | 1 | 1 | 701 | 1 | 7 |
| **Gleason Score** | | | | | | | | | |
| ≤6 | 42 | - | - | 63** | 23 | 4 | 132 | 1 | - |
| 7 | 99 | - | - | 271 | 16 | 16 | 402 | 132 | - |
| 8 | 52 | - | - | 68 | 8 | 3 | 131 | 26 | - |
| 9 | 54 | - | - | 134 | 1 | 5 | 194 | 77 | - |
| 10 | 1 | - | - | 9 | 0 | 1 | 11 | 0 | - |
| *NA* | 0 | 44 | 85 | 0 | 0 | 1 | 130 | 3 | 493 |
| **Preoperative PSA (ng/ml)** | | | | | | | | | |
| <10 | 50 | - | - | - | 21 | 15 | 86 | 123 | - |
| 10-20 | 95 | - | - | - | 17 | 9 | 121 | 92 | - |
| >20 | 103 | - | - | - | 8 | 6 | 117 | 24 | - |
| *NA* | 0 | 44 | 85 | 545 | 2 | 0 | 676 | 0 | 493 |
| **Metastasis** | | | | | | | | | |
| No | 226 | 36 | 34 | 333 | 39 | 26 | 694 | 146 | - |
| Yes | 22 | 8 | 51 | 212 | 9 | 4 | 306 | 93 | - |
| *NA* | 0 | 0 | 0 | 0 | 0 | 0 | 0 | 0 | 493 |

*Progression free-survival, overall survival not available, metastasis free-survival not available

### Overall-survival and metastasis free-survival is available, Progression-free survival is not available.

§ Progression-free survival and overall survival is available, metastasis free-survival not available. See text for more details.

** Gleason score 5 is present in both; 3/63 patients (4.7%) from GSE46691.

| | effectSize | effectSizeS | effectSizeF | effectSizeF | tauSquared | numStudie | cochranes | heterogene | fisherStatU |
|---|---|---|---|---|---|---|---|---|---|
| TMSB10 | 0.428054 | 0.073853 | 6.79E-09 | 7.2E-05 | 0 | 6 | 3.058317 | 0.690995 | 58.36341 |
| ENSA | 0.406589 | 0.072319 | 1.89E-08 | 9.21E-05 | 0 | 5 | 1.901073 | 0.753948 | 50.10471 |
| ASPN | 0.416526 | 0.076852 | 5.97E-08 | 0.000194 | 0.00028 | 4 | 3.021495 | 0.388323 | 47.87951 |
| YWHAZ | 0.38574 | 0.072989 | 1.26E-07 | 0.000351 | 0 | 6 | 0.234439 | 0.998698 | 46.67544 |
| HES6 | 0.340692 | 0.073513 | 3.58E-06 | 0.005419 | 0 | 6 | 2.889092 | 0.71708 | 42.04177 |
| STC2 | 0.325374 | 0.073489 | 9.53E-06 | 0.010948 | 0 | 6 | 4.894552 | 0.428884 | 44.73413 |
| ASNS | 0.315806 | 0.072853 | 1.46E-05 | 0.015824 | 0 | 6 | 4.766721 | 0.445008 | 39.32488 |
| HAVCR2 | 0.316027 | 0.073508 | 1.71E-05 | 0.016732 | 0 | 6 | 4.339751 | 0.501606 | 41.25217 |
| ARL6IP1 | 0.312278 | 0.072892 | 1.83E-05 | 0.017057 | 0 | 6 | 3.949728 | 0.556676 | 43.11247 |
| F5 | 0.312467 | 0.073472 | 2.11E-05 | 0.018247 | 0 | 6 | 3.769004 | 0.583129 | 38.72005 |
| RFTN1 | 0.312514 | 0.073554 | 2.15E-05 | 0.018247 | 0 | 6 | 4.941401 | 0.423073 | 40.68916 |
| SOX4 | 0.311313 | 0.073489 | 2.27E-05 | 0.018495 | 0 | 6 | 3.109998 | 0.683032 | 39.16073 |
| PTPN9 | 0.315421 | 0.074876 | 2.52E-05 | 0.019718 | 0 | 5 | 3.847943 | 0.426975 | 35.34021 |
| ALDH1A1 | 0.30538 | 0.072822 | 2.75E-05 | 0.019912 | 0 | 6 | 3.563329 | 0.613828 | 37.5135 |
| MRPL11 | 0.307843 | 0.073419 | 2.75E-05 | 0.019912 | 0 | 5 | 0.547532 | 0.968712 | 34.33387 |
| GABRD | 0.307954 | 0.075551 | 4.58E-05 | 0.027535 | 0 | 4 | 1.257899 | 0.739154 | 30.92954 |
| RC3H2 | 0.299169 | 0.073463 | 4.65E-05 | 0.027535 | 0 | 6 | 3.40777 | 0.637387 | 37.74427 |
| CST2 | 0.337784 | 0.084571 | 6.49E-05 | 0.037297 | 0.003776 | 6 | 5.346692 | 0.37505 | 44.95804 |
| CXCR4 | 0.539147 | 0.137466 | 8.78E-05 | 0.043732 | 0.048262 | 6 | 9.581595 | 0.087997 | 59.79677 |
| SEM1 | 0.28409 | 0.072802 | 9.53E-05 | 0.043732 | 0 | 6 | 4.402457 | 0.493039 | 35.13656 |
| FOXH1 | 0.293279 | 0.075438 | 0.000101 | 0.043732 | 0 | 4 | 0.42456 | 0.935124 | 28.4786 |
| KIF7 | 0.308666 | 0.079597 | 0.000105 | 0.043732 | 0.001942 | 6 | 5.17683 | 0.394683 | 42.53346 |
| BARD1 | 0.28314 | 0.073434 | 0.000115 | 0.043732 | 0 | 6 | 2.923426 | 0.71179 | 35.16521 |
| CADPS | 0.483603 | 0.126005 | 0.000124 | 0.043732 | 0.032064 | 6 | 7.893648 | 0.162195 | 60.59997 |
| RNF19A | 0.284064 | 0.074105 | 0.000126 | 0.043732 | 0 | 5 | 3.221527 | 0.521461 | 32.67187 |
| CAMK2N1 | 0.588686 | 0.155414 | 0.000152 | 0.047094 | 0.068719 | 6 | 11.163 | 0.048242 | 102.4518 |
| GPR37 | 0.276855 | 0.073392 | 0.000162 | 0.047862 | 0 | 6 | 4.189835 | 0.522421 | 37.15347 |

| fisherPvalU | fisherFDRU | fisherStatD | fisherPvalD | fisherFDRDown |
|---|---|---|---|---|
| 4.48E-08 | 8.75E-05 | 0.79757 | 0.999996 | 1 |
| 2.55E-07 | 0.000249 | 2.237338 | 0.994176 | 1 |
| 1.04E-07 | 0.000127 | 0.855222 | 0.999008 | 1 |
| 5.31E-06 | 0.002158 | 1.535464 | 0.999852 | 1 |
| 3.28E-05 | 0.00831 | 3.096242 | 0.994826 | 1 |
| 1.14E-05 | 0.003724 | 1.254979 | 0.99995 | 1 |
| 9.3E-05 | 0.018346 | 3.725536 | 0.987895 | 1 |
| 4.45E-05 | 0.010096 | 2.524479 | 0.998071 | 1 |
| 2.16E-05 | 0.006298 | 4.912793 | 0.960828 | 1 |
| 0.000117 | 0.02084 | 3.387598 | 0.992134 | 1 |
| 5.52E-05 | 0.012115 | 2.748858 | 0.997072 | 1 |
| 9.9E-05 | 0.018898 | 2.297544 | 0.998795 | 1 |
| 0.000109 | 0.020507 | 3.088226 | 0.979271 | 1 |
| 0.000184 | 0.027066 | 4.608721 | 0.969789 | 1 |
| 0.000162 | 0.025325 | 4.361036 | 0.929594 | 1 |
| 0.000145 | 0.023003 | 1.118132 | 0.997386 | 1 |
| 0.000169 | 0.025993 | 2.510651 | 0.998123 | 1 |
| 1.05E-05 | 0.003467 | 3.820648 | 0.98646 | 1 |
| 2.46E-08 | 5.33E-05 | 0.376627 | 1 | 1 |
| 0.000445 | 0.043977 | 7.760874 | 0.803527 | 1 |
| 0.000391 | 0.040858 | 0.93278 | 0.998638 | 1 |
| 2.71E-05 | 0.007243 | 0.772517 | 0.999997 | 1 |
| 0.000441 | 0.04391 | 2.556609 | 0.997947 | 1 |
| 1.75E-08 | 4.56E-05 | 0.319446 | 1 | 1 |
| 0.000309 | 0.035515 | 4.167829 | 0.939458 | 1 |
| 1.84E-16 | 3.59E-12 | 1.080587 | 0.999978 | 1 |
| 0.000211 | 0.029087 | 1.941948 | 0.99949 | 1 |

| | effectSize | effectSizeS | effectSizeF | effectSizeF | tauSquare | numStudie | cochranes | heterogene | fisherStatU |
|---|---|---|---|---|---|---|---|---|---|
| KCTD14 | -0.42649 | 0.073761 | 7.38E-09 | 7.2E-05 | 0 | 6 | 3.810372 | 0.577028 | 1.000633 |
| AZGP1 | -0.74251 | 0.131525 | 1.65E-08 | 9.21E-05 | 0.037163 | 6 | 8.268514 | 0.142045 | 0.086328 |
| PART1 | -0.41007 | 0.07383 | 2.79E-08 | 0.000109 | 0 | 6 | 4.959765 | 0.42081 | 0.805702 |
| CHRNA2 | -0.46132 | 0.092096 | 5.47E-07 | 0.001334 | 0.007013 | 6 | 5.631412 | 0.343751 | 0.420963 |
| DPT | -0.36083 | 0.073597 | 9.45E-07 | 0.001845 | 0 | 6 | 3.569939 | 0.612832 | 1.486749 |
| EDN3 | -0.34768 | 0.074149 | 2.75E-06 | 0.004875 | 0 | 5 | 3.883093 | 0.422059 | 2.233469 |
| KIAA1210 | -0.34707 | 0.075022 | 3.72E-06 | 0.005419 | 0 | 5 | 2.74928 | 0.600619 | 0.48043 |
| LTF | -0.33859 | 0.073572 | 4.18E-06 | 0.005419 | 0 | 6 | 4.66407 | 0.458236 | 0.508951 |
| SIDT1 | -0.31703 | 0.073461 | 1.59E-05 | 0.016357 | 0 | 6 | 4.504791 | 0.479243 | 1.35713 |
| CBLL1 | -0.30333 | 0.072806 | 3.1E-05 | 0.021587 | 0 | 6 | 3.182558 | 0.671864 | 3.823916 |
| PTN | -0.30287 | 0.073456 | 3.74E-05 | 0.025166 | 0 | 6 | 3.512919 | 0.621434 | 1.121276 |
| CCK | -0.29943 | 0.073426 | 4.54E-05 | 0.027535 | 0 | 6 | 2.232442 | 0.816136 | 2.532233 |
| UFM1 | -0.28275 | 0.07279 | 0.000103 | 0.043732 | 0 | 6 | 1.948897 | 0.85617 | 4.824567 |
| CPA3 | -0.28432 | 0.073459 | 0.000109 | 0.043732 | 0 | 6 | 2.604775 | 0.76064 | 1.873589 |
| CDC42EP5 | -0.60195 | 0.157306 | 0.00013 | 0.043732 | 0.071052 | 6 | 11.23511 | 0.046913 | 1.795606 |
| TMEM121B | -0.27885 | 0.073375 | 0.000144 | 0.046736 | 0 | 6 | 4.448048 | 0.486864 | 1.223399 |
| AKAP7 | -0.27848 | 0.073326 | 0.000146 | 0.046736 | 0 | 5 | 3.035462 | 0.551909 | 5.246321 |
| KLF4 | -0.27499 | 0.072762 | 0.000157 | 0.047249 | 0 | 6 | 4.529215 | 0.475984 | 2.333823 |

| fisherPvalU | fisherFDRU | fisherStatD | fisherPvalD | fisherFDRDown |
|---|---|---|---|---|
| 0.999986 | 1 | 58.87393 | 3.62E-08 | 0.000118 |
| 1 | 1 | 101.4587 | 2.88E-16 | 5.63E-12 |
| 0.999996 | 1 | 57.69179 | 5.93E-08 | 0.000165 |
| 1 | 1 | 60.73888 | 1.65E-08 | 8.08E-05 |
| 0.999876 | 1 | 47.46301 | 3.87E-06 | 0.003876 |
| 0.994217 | 1 | 40.0209 | 1.68E-05 | 0.008633 |
| 0.999995 | 1 | 42.13369 | 7.1E-06 | 0.0047 |
| 1 | 1 | 47.4022 | 3.97E-06 | 0.003876 |
| 0.999924 | 1 | 43.09723 | 2.17E-05 | 0.010612 |
| 0.986408 | 1 | 38.71555 | 0.000117 | 0.026926 |
| 0.999973 | 1 | 40.44399 | 6.07E-05 | 0.017949 |
| 0.998042 | 1 | 36.64045 | 0.000255 | 0.044535 |
| 0.963583 | 1 | 38.54051 | 0.000125 | 0.028285 |
| 0.999576 | 1 | 35.88091 | 0.000339 | 0.049337 |
| 0.999661 | 1 | 76.46852 | 1.94E-11 | 1.89E-07 |
| 0.999957 | 1 | 38.38424 | 0.000133 | 0.028823 |
| 0.874128 | 0.987361 | 32.80762 | 0.000293 | 0.047404 |
| 0.998696 | 1 | 38.1013 | 0.000148 | 0.031372 |
